# Using twin-pairs to assess potential bias in polygenic prediction of externalising behaviours across development

**DOI:** 10.1101/2023.12.13.23299910

**Authors:** Joanna K. Bright, Christopher Rayner, Ze Freeman, Helena M.S. Zavos, Yasmin I. Ahmadzadeh, Essi Viding, Tom A. McAdams

## Abstract

Prediction from polygenic scores may be confounded sources of passive gene-environment correlation (rGE; e.g. population stratification, assortative mating, and environmentally mediated effects of parental genotype on child phenotype). Using genomic data from 10,000 twin pairs, we asked whether polygenic scores from the recent externalising genome-wide association study predicted conduct problems, ADHD symptomology and callous-unemotional traits, and whether these predictions are biased by rGE. We ran regression models including within-family and between-family polygenic scores, to separate the direct genetic influence on a trait from environmental influences that correlate with genes (indirect genetic effects). Findings suggested that this externalising polygenic score is a good index of direct genetic influence on conduct and ADHD-related symptoms across development, with minimal bias from rGE, although the polygenic score predicted less variance in CU traits. Post-hoc analyses showed some indirect genetic effects acting on a common factor indexing stability of conduct problems across time and contexts.

## Introduction

Common externalising phenotypes, including conduct problems and attention-deficit/hyperactivity disorder (ADHD), are associated with adverse outcomes such as low academic achievement, social isolation and substance use disorders^1,2^. More recently, callous-unemotional traits (CU) have been assessed concurrently and appear to index children with particularly severe behavioural problems^3,4^. Previous research has shown that conduct problems, ADHD and CU traits are moderately to highly heritable^5,6^, share common aetiological influences^7^ and can be predicted by polygenic scores extracted from genome-wide association studies (GWAS)^8,9^. Polygenic scores are increasingly used as tools to predict risk profiles^10^ and it is hoped that in the future, polygenic indices will have clinical utility^11^. They can also be used by researchers as instrumental variables in causal inference analyses^12,13^.

It is increasingly understood that polygenic scores for complex traits may not simply index a person’s genetic liability. Instead, as genetic and environmental risks correlate, polygenic scores may also capture sources of passive gene-environment correlation (rGE), including population stratification, assortative mating, and the indirect genetic effects of parental genotype on child phenotype via parental behaviour (genetic nurture)^14–19^. Without accounting for such rGE, we cannot be sure of the true relationship between individuals’ direct genetic liability and subsequent behaviour.

Research has repeatedly shown that associations between polygenic scores and cognitive traits/ educational attainment are biased by rGE^15,19,20^. Symptoms related to conduct problems, ADHD, and CU traits have many features which make it plausible that polygenic prediction may also capture more than the direct genetic effects of a person’s genome on their phenotype. Previous research has suggested considerable assortative mating for externalising behaviours^21,22^ and the family environment is hypothesized to be a key contributor to externalising symptoms in children, so genetic nurture is a possibility^16^. Furthermore, externalising behaviours are strongly associated with socioeconomic status (SES) and educational attainment^23,24^, which are subject to population stratification, assortative mating, and parental effects^25^. Family-based models of externalising features have suggested some bias from rGE acting on externalising phenotypes. For example, associations between externalising polygenic scores and child ADHD are attenuated when parents’ genetic liability transmitted via the rearing environment was accounted for^26^. Similarly, single nucleotide polymorphism (SNP) heritability estimates for conduct disorder and ADHD symptoms were attenuated after accounting for the effects of parental genotype^16^. Findings are not consistently replicated across studies, however, with some reporting no evidence for indirect genetic effects on ADHD phenotypes^19,27^. However, these studies reporting null findings used polygenic scores from an ADHD GWAS of a smaller sample than that used for the most recent externalising GWAS^28^, and predicted into samples smaller than those that found evidence for indirect genetic effects^16,26^. As GWASs become larger, the variance explained by resultant polygenic scores tend to increase^29^. Consequently, there is more statistical power to detect portions of the variance attributable to passive rGE, not just direct genetic effects.

The potential benefit of using polygenic scores to elucidate variation in human behaviour is undermined if we do not systematically examine possible biases introduced by rGE. Using sibling pairs in genomic analyses allows estimation of direct genetic effects free from such biases, as siblings are matched for family environment (i.e. they share the effects of population stratification, assortative mating, and genetic nurture)^14^. Dizygotic (DZ) twin pairs are additionally matched for prenatal environment and time-variant factors. By using DZ twin pairs in our genomic analyses, we can thus account for some of the potential biases in polygenic prediction of our phenotypes of interest and get closer to capturing true effects of direct genetic influences within individuals. In short, by comparing polygenic prediction of phenotypes within-families (i.e., comparing prediction between DZ twins within a family) and between families (comparing prediction between family units in a sample), we can separate the direct genetic influence on a trait from environmental influences that correlate with genes.

Here, we combined genomic and family data to investigate polygenic prediction for conduct problems, ADHD symptomology and CU traits, and the degree to which these predictions may be biased by rGE. Our approach was developmental, focusing on multiple timepoints from ages 4-21 years, looking across parent, child, and teacher reports. We used a polygenic score from the most recent externalising GWAS of one million individuals of European ancestry^8^. Where we found evidence for bias from indirect genetic effects in polygenic prediction, we tested whether measures of SES, neighbourhood deprivation, and parenting behaviours explained that bias, and whether these variables impacted estimates of direct genetic effects. We complemented our polygenic analyses of DZ twins with univariate twin analyses including both monozygotic (MZ) and DZ twins^30^. It has been suggested that genetic nurture effects may be captured in twin study estimates of the shared environment, as they are shared between twins and should promote similarity among both MZ and DZ twins^18,27,31^. We therefore assumed that the magnitude of shared environmental estimates derived from twin analyses would be informative regarding the presence or absence of indirect genetic effects.

Our study addressed four key questions that have not been examined before. First, to what extent does the new polygenic score derived from a large-scale study of externalising-related behaviours index genetic liability for conduct problems, ADHD symptoms and CU traits respectively? Second, do the associations between the externalising polygenic score and these traits partly reflect environmental biases arising from sources of rGE? Third, does the degree of prediction and/or biases vary across development? Fourth, do environmental effects found in univariate twin models predicted presence of indirect genetic effects in genomic analyses? We hypothesised that the degree of prediction from the polygenic score to our phenotypes would be modest, in line with the currently observed magnitudes of variance accounted for by polygenic scores. We also hypothesised that prediction would be stronger for conduct problems and ADHD symptoms than for CU traits, given that prior research has indicated that CU traits have some level of genetic independence from broader externalising phenotypes^32–34^. Thirdly, we hypothesised that conduct problems would be most impacted by indirect genetic effects as prior research has typically reported higher estimates of shared environment for conduct problems than for ADHD symptoms or CU traits. We did not make specific hypotheses regarding the developmental effects.

## Results

### Cross-sectional polygenic analyses

The externalising polygenic score predicted 0.1-2.3% of the variance in conduct problems, ADHD symptoms and CU traits. We found a similar pattern of results for conduct disorder and ADHD symptomology (see Figure 1a and 2a). The polygenic score predicted an average of 1.4% of the variance in the phenotype for conduct problems and 1.3% for ADHD symptoms. There was a significant increase in prediction for parent-reported conduct problems over development, from 0.6% variance explained at age 4, to a peak of 1.9% at age 16. Similarly, prediction rose from 0.5% to 1.8% of the variance in parent-rated ADHD symptoms from age 4 to 21. At each timepoint, there was no significant difference in polygenic prediction between reporters for conduct problems or ADHD symptoms. Polygenic prediction of conduct problems and ADHD symptoms was almost wholly due to direct genetic effects at all ages, i.e. there was no evidence for a role of indirect genetic effects on these externalising outcomes. The only exception to this was for teacher-reported conduct problems at age 9, we did find significant indirect genetic effects (Figure 1a).

**Figure 1:**
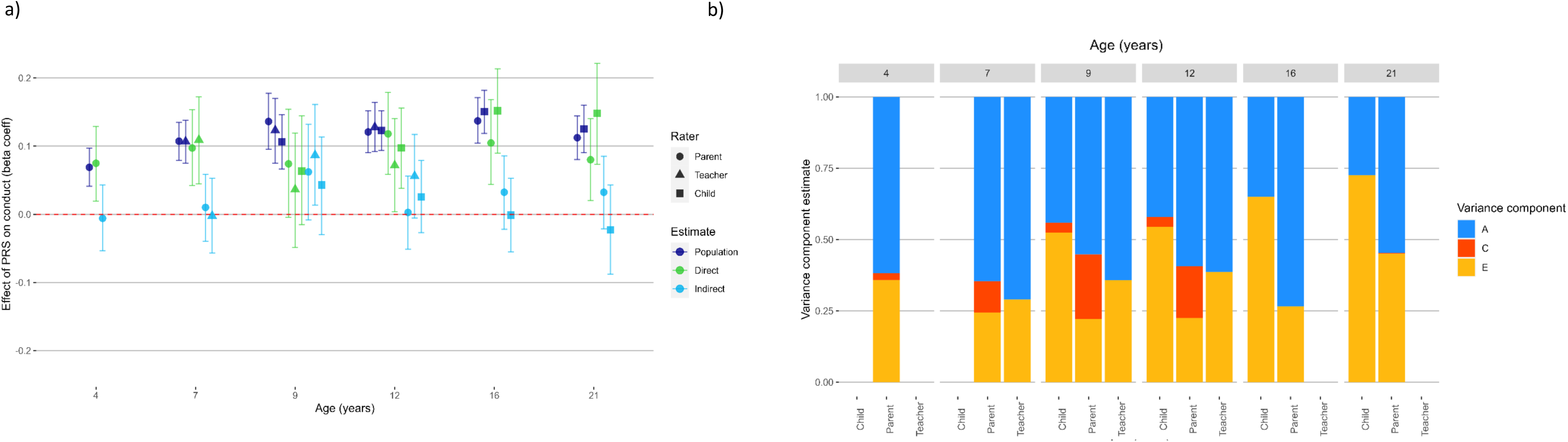
a) Estimating population-level, direct and indirect genetic effects of externalising PRS and b) univariate ACE twin models for conduct problems. Beta coefficient estimates of population-level prediction of externalising PRS for conduct problems, alongside estimates of direct and indirect genetic effects, reported across developmental timepoints and reporter. Estimates of the contribution of additive genetic effects (A), common environmental influences (C) and unique environmental influences (E) in the variance of conduct problems. Each univariate twin model was repeated at each timepoint, and for parent, teacher, and self-reports.

**Figure 2:**
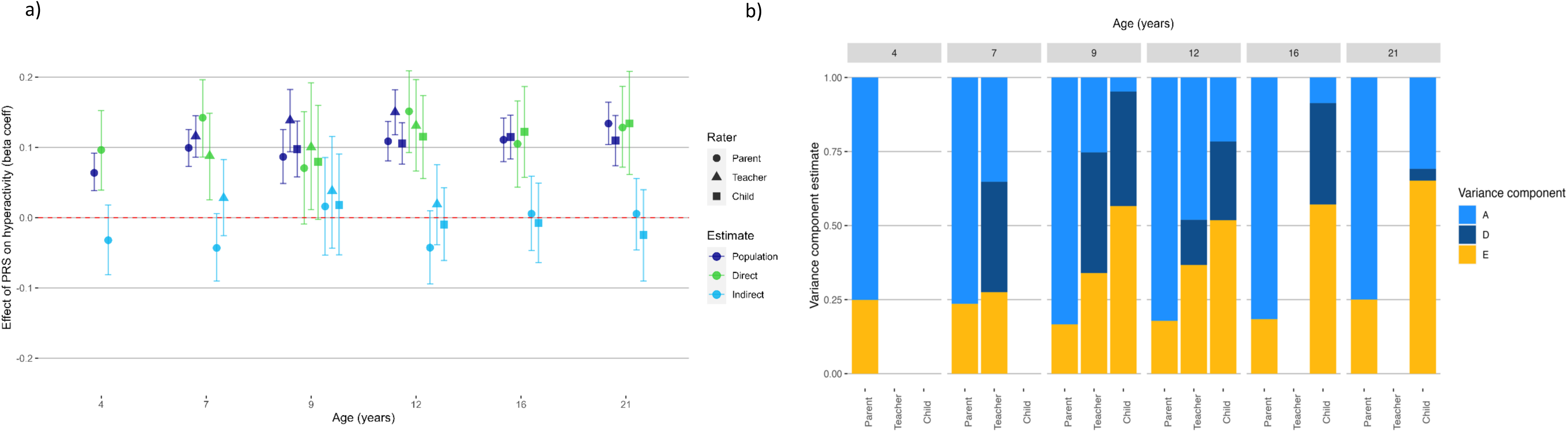
Estimating a) population-level, direct and indirect genetic effects of externalising PRS and b) univariate ADE twin models for ADHD symptoms across development and reporter. Beta coefficient estimates of population-level prediction of externalising PRS for hyperactivity problems, alongside estimates of direct and indirect genetic effects, reported across developmental timepoints and reporter. Estimates of the contribution of additive genetic effects (A), dominant genetic effects (D) and unique environmental influences (E) in the variance of hyperactivity problems. Each univariate twin model was repeated at each timepoint, and for parent, teacher, and self-reports. D was dropped from the parent models as it was not significant once sibling interaction terms were included.

The polygenic score predicted less of the variance in CU traits, averaging 0.4% of variance explained (Figure 3a). There was a significant difference between the polygenic prediction of parent and teacher reported CU traits at age 9. There was no change in magnitude of prediction between ages 7 and 12, although at age 16 the polygenic score had a negative association with parent-reported CU traits. We did not find significant direct or indirect genetic effects on CU traits once we broke the polygenic prediction down into within-family and between-family effects.

**Figure 3:**
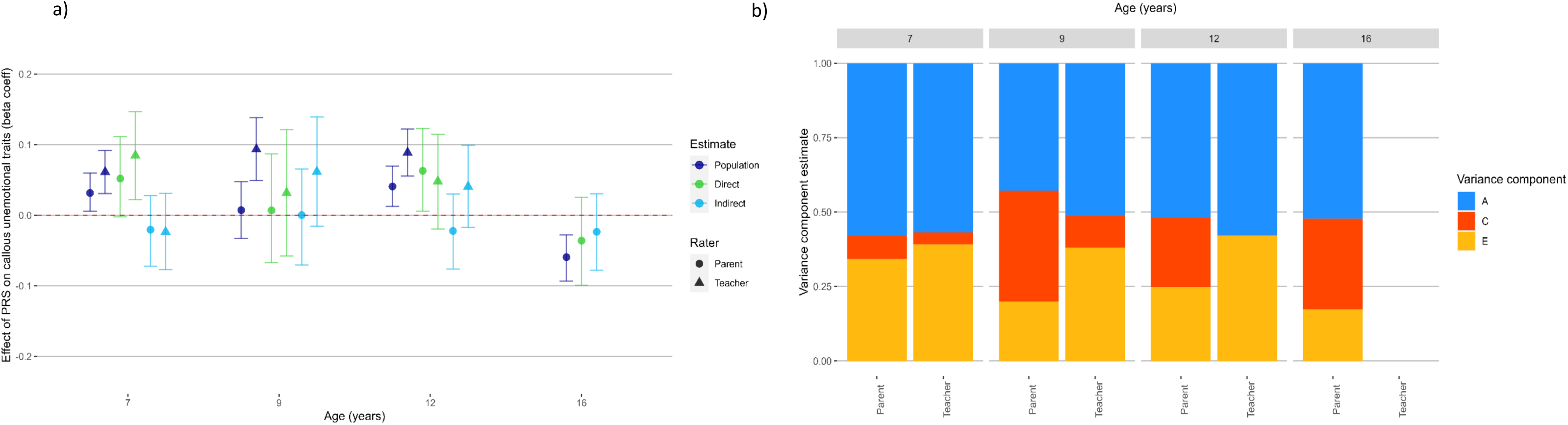
Estimating a) population-level, direct and indirect genetic effects of externalising PRS and b) univariate ACE twin models for callous-unemotional traits across development and reporter. Beta coefficient estimates of population-level prediction of externalising PRS for an index of callous-unemotional traits, alongside estimates of direct and indirect genetic effects, reported across developmental timepoints and reporter. Estimates of the contribution of additive genetic effects (A), common environmental influences (C) and unique environmental influences (E) in the variance in an index of callous-unemotional traits. Each univariate twin model was repeated at each timepoint, and for parent and teacher reports.

### Cross-sectional twin analyses

The results from twin models are shown in Figures 1b, 2b and 3b. Shared environmental effects (C) were estimated as significant for parent reported conduct problems at age 7 (11% of variance), age 9 (23%) and age 12 (18%). Model fit for ADHD symptoms was best when dropping C for D (dominant genetic effects; see Supplementary Materials). For CU traits, estimates of C were significant for parent reported CU at all ages: explaining between 7% - 37% of variance. C also significantly explained 11% of the variance in teacher-reported CU traits at age 9. However, in polygenic analyses significant indirect genetic effects were not detected for any of these variables, suggesting that significant shared environment estimates were not predictive of indirect genetic effects. See Supplementary Materials for a more detailed discussion of the results from twin models.

### Examining potential sources of indirect genetic effects

As we found indirect genetic effects acting on polygenic prediction of teacher-reported conduct problems at age 9, we re-ran these models controlling for SES, neighbourhood deprivation or parenting-related factors to assess whether any potential indirect genetic effects were captured by these covariates (Figure 4). Estimates of direct genetic effects were minimally impacted by including these covariates. Further, we found that controlling for either neighbourhood deprivation or parenting behaviours entirely accounted for removed the indirect effect (Figure 4; 95% confidence intervals included zero). When controlling for SES, the 95% confidence interval for the indirect genetic effect did not include zero, although the lower interval was very close, at 0.003 (Figure 4).

**Figure 4:**
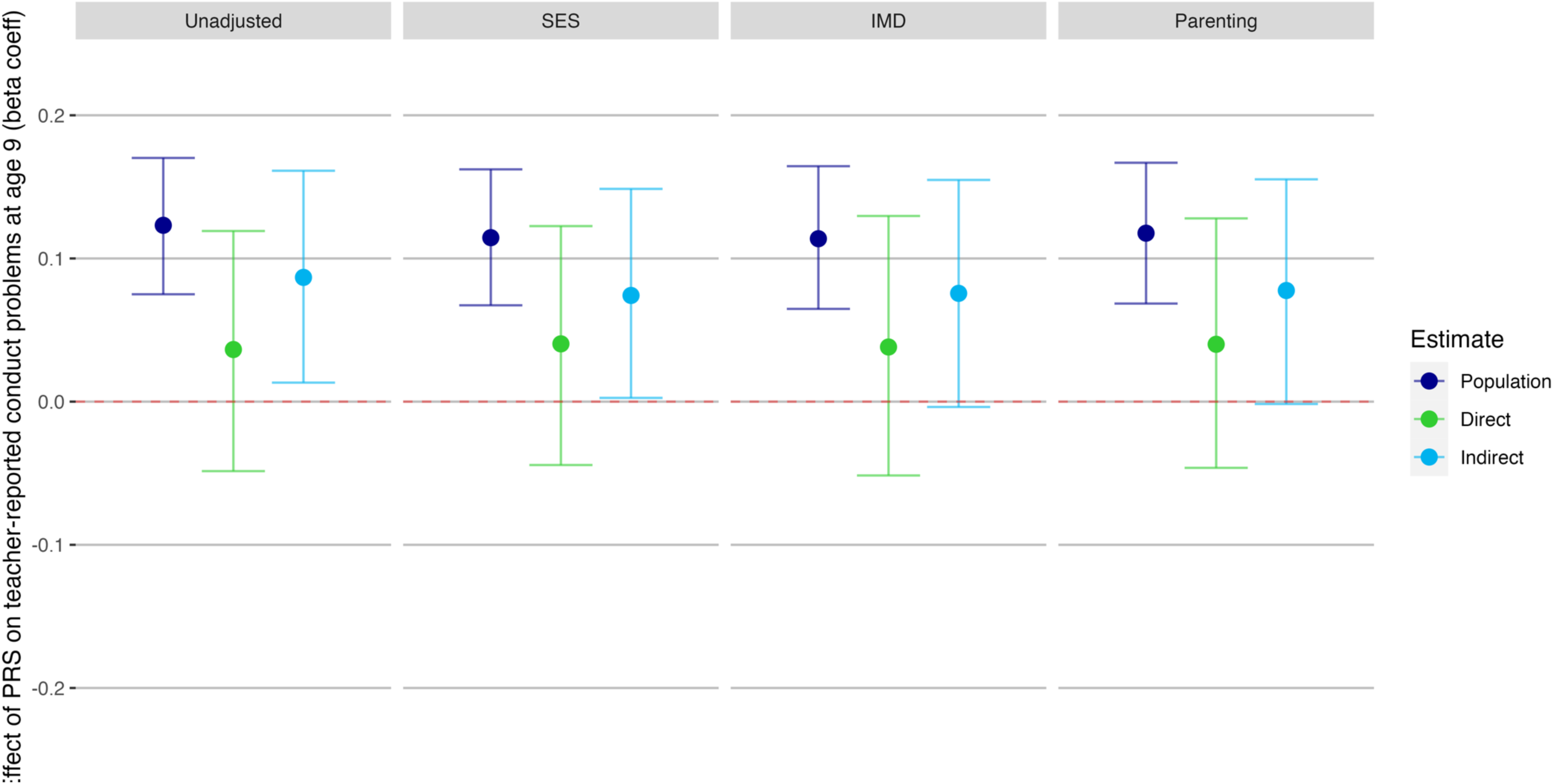
Investigating impact of socioeconomic status, neighbourhood deprivation or parenting variables on direct and indirect genetic effects of externalising PRS on teacher-reported conduct problems at age 9. Beta coefficient estimates of population-level prediction of externalising PRS, alongside estimates of direct and indirect genetic effects, for teacher-reported conduct problems at age 9, where we found significant indirect genetic effects. Socioeconomic status (SES) was measured at first contact and comprises measures of parent employment, education, and age of mother on first birth. The Indices of Multiple Deprivation (IMD) decile score uses census data matched with participants post codes, giving a broader measure of wider environmental factors such as local levels of employment and education, crime rates, barriers to housing and living environment quality. The parenting analyses used a latent factor created from ‘Parental Feelings’ and ‘Parental Discipline’ rated by parent at ages 4, 7, 9 and 12.

### Post-hoc common factor analyses: stability across time and contexts

We extracted common factor scores for each phenotype to index trait stability across time and reporter (Figure 5a). The polygenic score predicted 3.4% of variance in the conduct problems factor, 2.9% of variance in the ADHD symptoms factor and 1.0% of variance in the CU traits factor. Using these factors in the within- and between-family regression models, we found significant indirect genetic effects on conduct problems, which made up 40% of the total prediction. For ADHD symptoms and callous-unemotional traits, the prediction was wholly due to direct genetic effects. Further analyses showed that these results were consistent when stratified by reporter or timepoint (Supplementary Figures 4a & 5a). When using a common factor score for callous-unemotional traits, we were able to detect some population and direct genetic effects, whilst indirect genetic effects remained non-significant.

**Figure 5:**
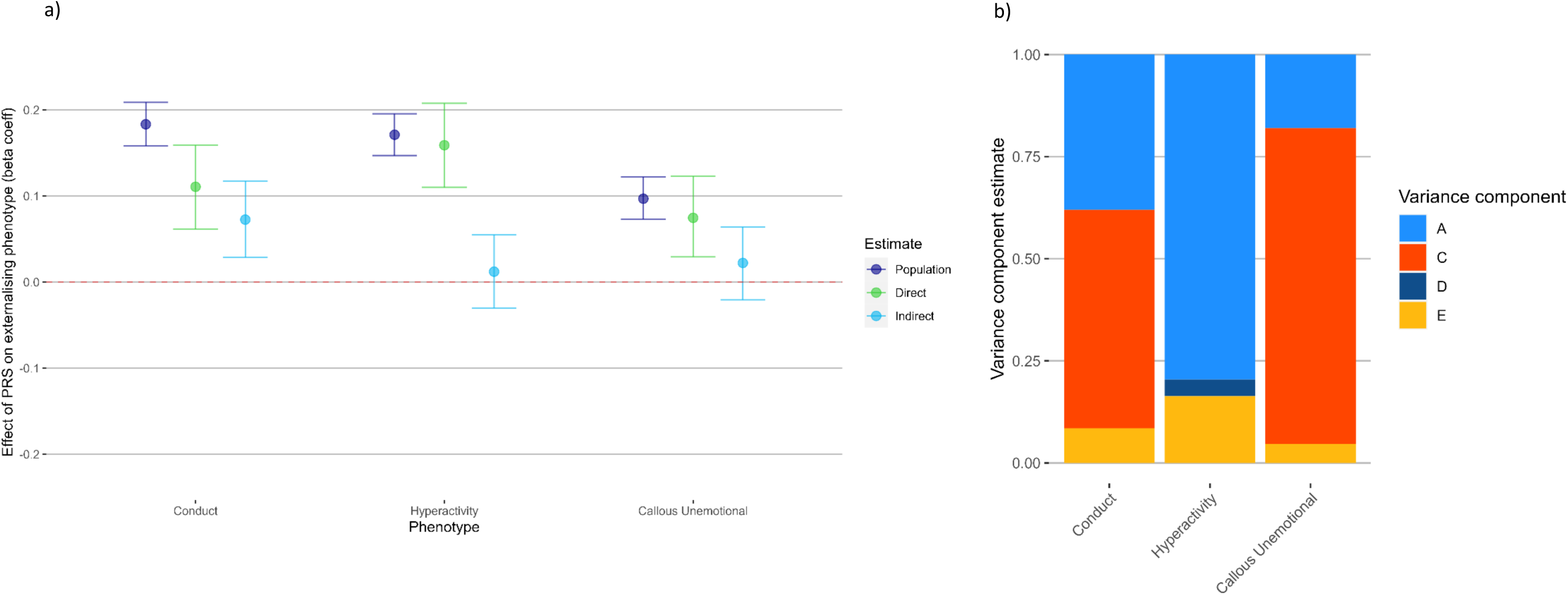
Estimating a) population-level, direct and indirect genetic effects of externalising PRS and b) univariate ACE twin models for common factor scores for each phenotype. Factor scores were created using common factor analysis in lavaan, extracting stability for each phenotype from measures across all timepoints and reporter. Beta coefficient estimates of population-level prediction of externalising PRS alongside estimates of direct and indirect genetic effects, for common factor scores created for conduct problems, ADHD symptoms and callous-unemotional traits. Estimates of the contribution of additive genetic effects (A), common environmental influences (C) and unique environmental influences (E) in the variance of each factor.

Where we found indirect genetic effects acting on polygenic prediction of stability (in conduct problems), we re-ran these models controlling for SES, neighbourhood deprivation or parenting-related factors (Figure 6). Estimates of direct genetic effects were minimally impacted by including these covariates (attenuation of between 2-11% of the effect sizes found in uncontrolled analyses). For the common factor score of conduct problems, we found that controlling for SES reduced the indirect genetic effect to zero (Figure 6). Neither the index of neighbourhood deprivation nor parenting behaviours impacted the significance of the indirect genetic effects on the common factor score for conduct problems, however they did account for the indirect genetic effect on two out of four within-time factors. Controlling for parenting led to a non-significant effect of the polygenic score on within-twin and within-teacher factors.

**Figure 6:**
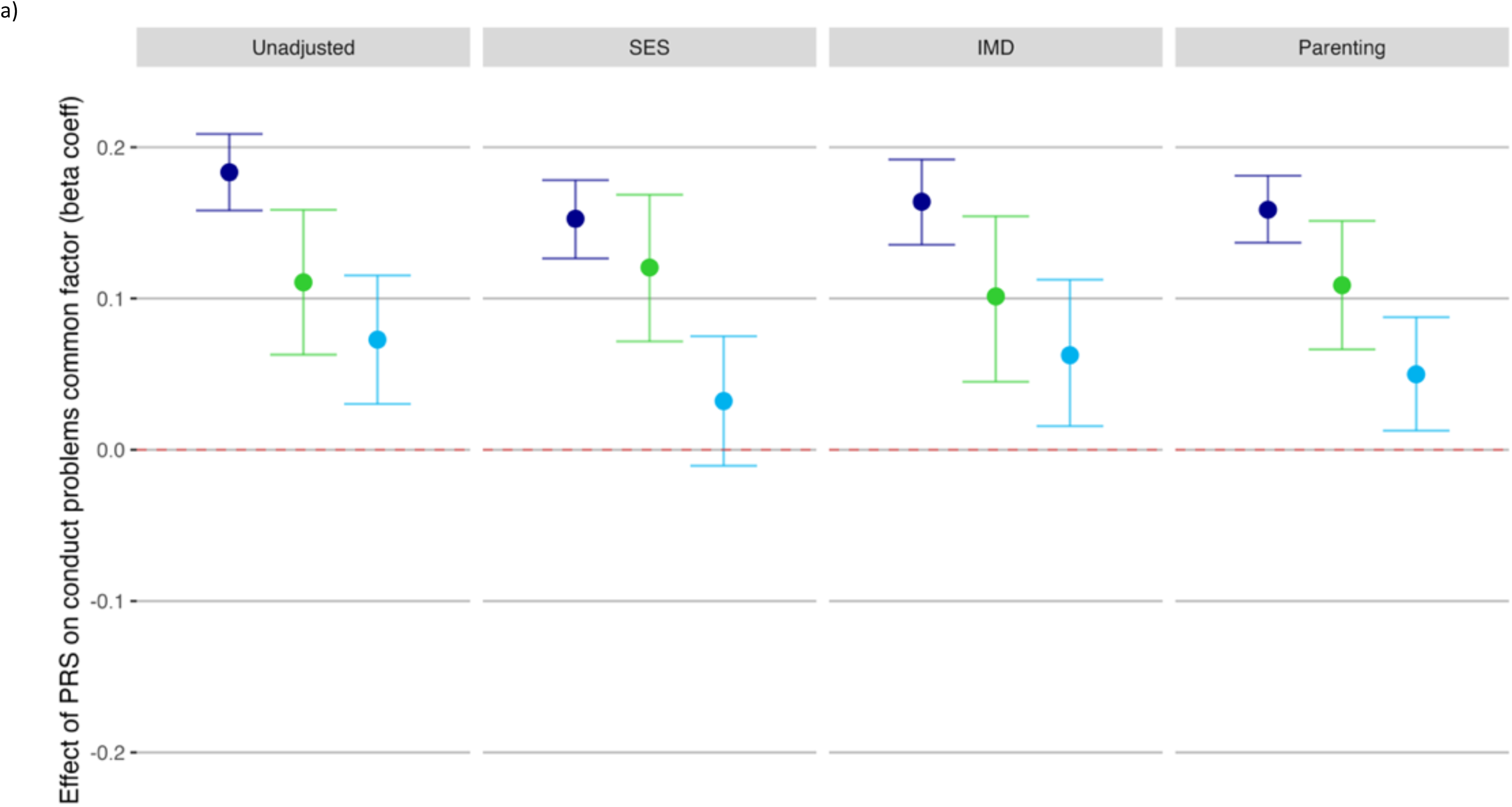
Investigating impact of socioeconomic status indices or parenting variables on direct and indirect genetic effects of externalising PRS on the common factor score for conduct problems. Socioeconomic status (SES) was measured at first contact and comprises measures of parent employment, education, and age of mother on first birth. The Indices of Multiple Deprivation (IMD) decile score uses census data matched with participants post codes, giving a broader measure of wider environmental factors such as local levels of employment and education, crime rates, barriers to housing and living environment quality. The parenting analyses used a latent factor created from ‘Parental Feelings’ and ‘Parental Discipline’ rated by parent at ages 4, 7, 9 and 12.

When using a common factor score in twin models (Figure 5b), we found a much larger presence of C for conduct problems (80%) and CU traits (77%) than in the cross-sectional analyses. When we broke down each factor into within-reporter factors (Supplementary Figures 4b and 5b), we can see that this large estimate of C was driven by parent-reports, for both conduct problems and CU traits. For ADHD symptoms, A accounted for the majority of variance, with minimal influence of D or E. Although significant C estimates in twin analyses did not reliably predict the presence of indirect genetic effects, where indirect genetic effects were detected in polygenic score analyses (i.e. for conduct problems), C was consistently significant in twin analyses.

## Discussion

Polygenic scores hold great potential for better understanding the developmental aetiology of psychiatric and behavioural problems, including externalising phenotypes. However, studies that examine associations between polygenic scores and such outcomes should also investigate possible sources of passive gene-environment correlation (rGE). Passive rGE may confound associations between individuals’ direct genetic liability and subsequent behaviour, so before we conclude that polygenic scores represent direct genetic risk, this assumption should be examined. We tested for indirect genetic confounding in associations between a polygenic score for externalising (derived from a GWAS of over one million people of European Ancestry)^8^ and parent-, teacher- and self-reported measures of conduct problems, ADHD symptoms, and callous-unemotional (CU) traits by using dizygotic twin pairs from a developmental twin cohort. Findings from our main, pre-registered analyses suggested that this externalising polygenic score is a good index of direct genetic influence on conduct and ADHD-related symptoms across development, with minimal bias from rGE. For CU traits, the polygenic score predicted less variance.

In cross-sectional analyses we found no statistically significant indirect genetic effects on either conduct problems or ADHD-related symptoms at any age or for any reporter., except teacher-reported conduct problems at age 9. This aligns with findings by authors of the externalising GWAS who found that whilst there was attenuation of the externalising polygenic prediction of a phenotypic externalising factor when using within family models, this prediction remained significant^8^. It is possible that the indirect genetic effects found on teacher-reported conduct problems at age 9 is a spurious finding: the TEDS sample at age 9 was half the size of other timepoints, with non-significant direct genetic effects. For parent and child reported conduct problems at age 9, neither direct nor indirect effects appeared significant, suggesting we lacked the power to accurately decompose the signal in the data available at this time point. Overall, in this sample of developing children, the externalising polygenic score served as a good marker of direct genetic influence on conduct problems and ADHD symptoms, seemingly unbiased by indirect genetic effects such as population stratification and genetic nurture.

The externalising polygenic score provided a much lower prediction of CU symptoms than conduct problems or ADHD symptoms. Given the externalising polygenic score used in this study was derived using phenotypes that were more closely related to conduct problems and ADHD than CU traits, it is perhaps not surprising that the prediction was less strong. Twin data also indicates only moderate overlap between heritability of conduct problems/ADHD symptoms and CU traits^32–34^. Therefore, our findings are line with the notion that although CU traits are associated and share genetic risk factors with conduct problems and ADHD symptoms, they are also influenced by genes not shared with these externalising phenotypes^7^.

When looking across development, from the ages 4 to 21 years, we found a small, significant increase in population-level polygenic prediction of conduct problems and ADHD symptoms. This may depict the genes involved in the presentation of these externalising symptoms having a stronger influence later in childhood and early adulthood. However, it is important to consider that the polygenic scores used here were created from a GWAS made from meta-analysed GWAS samples capturing a range of ages from children to much older adults. Previous research has shown that genetic influences on the intercept and slope of conduct problems, ADHD symptomatology, and CU traits are substantial, but largely non-overlapping^36–38^. This means the genes impacting the initial risk for developing these phenotypes and those impacting their developmental course appear to be at least partially distinct. Therefore, as our genetic index was not created specifically for young children, the prediction may not be equally as good in young children as compared to early adulthood, due to developmental genetic effects.

We had anticipated that associations between an externalising polygenic score and conduct problems, ADHD symptoms and CU traits could be subject to bias from rGE. However, our main analyses did not find evidence for this. One possible explanation for this may be that the genes picked up in the GWAS are not the same genes that drive rGE. The polygenic score accounted for a maximum of 2.3% of conduct problems, ADHD symptom and CU trait variance, which suggests there are genetic effects not captured in this polygenic score, which are yet to be found. We ran exploratory post-hoc analyses where we examined whether a different picture would emerge if we indexed stability on the externalising traits using common factor analyses across time and reporter. These yielded increased prediction from polygenic scores, and we also found a significant indirect genetic effect which accounted for 40% of the population-level prediction for conduct problems. This indirect genetic effect on conduct problems remained using within-reporter across-time and within-timepoint across-reporter common factors. These time and/or context stable indices capture something more trait-like than measures at a single time-point from a single reporter, reducing error and increasing statistical power. In our analyses, it seems that indirect genetic effects are present (or at least large enough to detect in the present sample) for conduct problems, but only when focussing on stable variance. Similarly, with the increased power of using a common factor score for CU traits, we were able to detect some direct genetic effects, whilst indirect genetic effects remained non-significant.

Research has suggested that stable sources of environmental influence on child behaviour such as SES and parenting behaviours may correlate with genetic risk and so contribute to indirect genetic effects^19^. Therefore, we ran analyses controlling for influences of SES, neighbourhood deprivation and parenting behaviours where we found significant indirect genetic effect; i.e. teacher-reported conduct problems at age 9 and common factor score analyses for conduct problems. For teacher-reported conduct problems at age 9, we found that controlling for neighbourhood deprivation or parenting behaviours accounted for the indirect effect. The indirect genetic effect on the common factor for conduct problems was no longer present once SES was included in the model, whilst the direct genetic effect showed minimal attenuation. This is encouraging for researchers who want to work with these polygenic scores to predict conduct problems but who do not have twin or family data, as our results suggest that including a measure of individual-level SES can account for bias from indirect genetic effects without impacting the estimation of direct genetic influence. It is notable that unlike conduct problems, we did not find indirect genetic effects for ADHD symptoms or CU traits, even when focussing on latent indices of stability. This was despite the conduct problems and ADHD symptoms having similar SNP heritability, suggesting similar power to detect indirect genetic effects in the ADHD analyses. This may point to differing influence of parent- and family-related factors on the stability of conduct problems and ADHD symptoms, aligning with prior published twin research which often describes shared environmental influences on conduct problems, but less often finds such influences for ADHD or CU traits^32,36,39^. The polygenic score predicted less variance in CU traits than conduct problems, so further investigation is needed with a more appropriate polygenic score for CU traits to determine whether we are capturing a true direct genetic effect or if there is an issue of power to detect indirect effects. Finally, we hypothesised that indirect genetic effects in the polygenic analysis may relate to estimates of shared environmental influence (C) in the twin models. We found very little evidence for indirect genetic effects, however for those indirect genetic effects that we did find, C was also present in the complementary twin analyses.

We note some limitations to our analyses. The sample at age 9 is half the size of other time-points, and this is where we found the only significant indirect genetic effect in the cross-sectional analysis. The mismatch in sample size limits comparison to other timepoints and interpretation of the significant finding. Secondly, the GWAS that was used to derive our polygenic scores includes phenotypes more strongly associated with conduct problems and ADHD symptoms. Therefore, the polygenic scores used may not be entirely suitable for predicting direct genetic liability for CU traits. Finally, for genomic analysis, the samples used in the GWAS were restricted to individuals with a European ancestry^8^, as were the individuals included in genotyping in TEDS (99.9% white)^40^. Thus, we cannot generalise these results to non-European ancestry populations. Future research should test this polygenic score in other samples to confirm whether these results replicate across populations.

This study broke new ground by taking advantage of family-based samples to systematically evaluate the predictive power of the latest externalising polygenic score in explaining variation in conduct problems, ADHD symptoms and CU traits across development. A particular strength of this study was the use of multiple study designs to draw inference regarding direct and indirect genetic effects. No prior study using polygenic scores has focused on these three phenotypes simultaneously and we took a novel developmental approach, using the same measures at multiple timepoints. We found robust evidence for direct genetic effects of the best-powered polygenic score for externalising on conduct problems and ADHD symptoms, that appeared consistent across reporters. We also demonstrated indirect genetic effects on the stability of conduct problems, which warrant further investigation. The externalising polygenic score predicted less variance in CU traits, suggesting a partially distinct, genetic aetiology for CU traits. Our study highlights the importance of considering the measures, constructs and analyses we use, as we seek to understand developmental risk for externalising problems and CU traits and to prevent these adverse outcomes.

## Methods

This study was preregistered on the Open Science Framework (https://osf.io/zh23d). Any additional non-preregistered analyses are considered exploratory and indicated in the text, with details in the Supplementary Materials.

### Sample

We used data from the Twins Early Development Study (TEDS; N∼=10,000 families)^41^, a cohort study of twins born in England and Wales between 1994 and 1997. We used available data from the whole sample when running univariate twin models, whereas for the genomic analyses we used a sub-sample of DZ pairs only (N=7,063 pairs, 99.9% white^40^). Data were used from waves collected when twins were 4, 7, 9, 12, 16 and 21-years-old, rated by their parents, their teachers and self-reported at later ages. Supplementary Table 1 shows sample size and reporters at each wave. NB the TEDS sample at age 9 was half the size of other timepoints due to funding and operational constraints. See Supplementary Materials for information on exclusion criteria and ethical approval.

#### Phenotypes

##### Conduct problems and ADHD symptoms

The Conduct Problems and Inattention-Hyperactivity subscales were taken from the Strengths and Difficulties Questionnaire (SDQ), a questionnaire aimed at identifying problem behaviours in children^42^. These were collected at twin ages 4, 7, 9, 12, 16 and 21 years of age.

##### Callous-unemotional traits

We created an index of callous-unemotional (CU) behaviours, from items collected at 7, 9, 12, and 16 years^38^. See Supplementary information for more information.

### Environmental Covariates

#### Socioeconomic status (SES)

We used the TEDS index of SES, measured at first contact. The index is a composite score created from the standardised average of measures of mother and father employment levels, mother and father educational levels, and mother’s age on birth of first child^43^.

#### Neighbourhood deprivation

The Index of Multiple Deprivation (IMD), is an index of neighbourhood deprivation created using participants’ post codes, giving a broader measure of wider environmental factors such as local levels of employment and education, crime rates, barriers to housing and living environment quality. More information on this score can be found on the UK government’s website (https://www.gov.uk/government/statistics/english-indices-of-deprivation-2010).

#### Parenting

We created a latent parenting factor from measures of parenting available at each timepoint in the TEDS dataset. These consisted of ‘Parental Feelings’ and ‘Parental Discipline’ rated by the registered primary parent at ages 4, 7, 9 and 12. The factor was created using the lavaan package in R.

### Polygenic scores

Polygenic scores for externalising were created for DZ twin pairs using summary statistics from the current most recent GWAS of externalising liability in individuals of European ancestry (N=1,045,957)^8^. Polygenic scores were computed using LDPred2^44^. For more information on polygenic score generation, see Supplementary Materials.

#### Analyses

For each phenotype and time point, we ran two complementary analyses: within/between family polygenic score analyses and univariate twin models. All data quality control and statistical analyses were conducted in R version 4.3.1^45^.

##### Polygenic score analyses

To estimate the contribution of direct and indirect genetic effects on externalising traits we ran two linear regression models in the DZ twin sample for each outcome (conduct problems, ADHD, and CU traits)^15^.

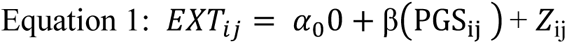

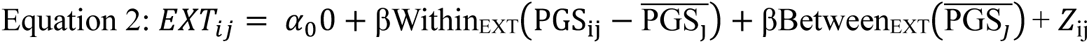

The first model estimated the population-level effect and included the polygenic score as a fixed effect (Equation 1, where for twin *i* in twin-pair *j,* EXT denotes the externalising outcome, and PGS is externalising polygenic score). The second model included the within-family and between-family polygenic scores as separate fixed effects (Equation 2)^15^. Here, the between family score is simply the family-based (twin-pair-based) mean polygenic score. The within-family polygenic score is the between family polygenic score subtracted from the individual’s polygenic score (twin *i*’s polygenic score minus their twin-pair’s mean score, 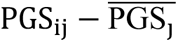) and between-family terms (the twin-pair’s polygenic score, 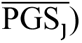. We included age, sex, age*sex, genotyping platform and the first 10 ancestry principal components in the Z term, as covariates in the models.

Extracted the effects of the polygenic scores from these regressions were used to calculate the indirect genetic effect by subtracting the direct genetic estimate (within-family effect) from the population estimate (between-family effect).

We computed bootstrapped standard errors and bias corrected confidence intervals for all effect estimates (population, direct and indirect), using the boot function, with 10,000 replications.

##### Examining potential sources of indirect genetic effects

Where indirect genetic effects were found, we ran analyses controlling for SES, neighbourhood deprivation or parenting behaviours accounted for these indirect genetic effects.

### Univariate twin models

We ran univariate twin models to evaluate whether derived aetiological estimates give any insight into whether indirect genetic effects may be present in associations between polygenic scores and our phenotypes. Specifically, it has been noted that indirect genetic effects share much in common with the shared environment, so we asked whether non-zero estimates of shared-environment influence predicted non-zero estimates of indirect genetic effect. Univariate twin models were applied to decompose phenotypic variation into additive genetic (A), dominant genetic (D) or shared environmental (C), and non-shared environmental (E) variance components, known as ADE or ACE models respectively (see Supplementary Figure 1)^30^. Analyses were run using the OpenMx R package^47^. All outcomes were adjusted for covariates (age, sex and sex*age). Contrast effects, where parents of non-identical twins contrast their twins and overestimate their differences, have been shown in parent-reported ADHD for dizygotic twins^46,48^. To control for this phenomenon, we included a sibling-interaction term into the univariate twin models for parent-reported ADHD symptoms.

#### Common factor analyses (not pre-registered)

After examining our data, we ran exploratory, post-hoc analyses using common factor scores for each phenotype computed from all available measures capturing stability across time and reporters. Although we did not pre-register these analyses, we believe these are important to include to contextualise our main findings, since genetic signal tends to be greater on indices of behavioural stability across time/contexts than on cross-sectional time/context specific measures, as common factor scores reduce measurement error, compared measures specific to a single timepoint or context^48,49^. We reasoned that running such analyses might further increase our power for detecting direct and indirect genetic effects. Common factor scores (Supplementary Figures 2-3) were computed using the cfa() function in the lavaan package for R^50^. We computed reporter- and age-specific factor scores to test whether effects varied by reporter or developmental stage. These additional results explore the extent to which focussing only on time/context specific analyses reduces our ability to detect genetic signal, and thus to distinguish direct from indirect genetic effects.

## Supplementary Methods

### Sample

Twins were excluded from analyses following standard TEDS protocols, which removes those with serious medical conditions which hinders ability to participate, those who experienced extreme adverse perinatal circumstances, and those with missing essential background variables, including sex/gender or zygosity (https://teds.ac.uk/datadictionary/exclusions.htm). The Twins Early Development Study (TEDS) was approved by King’s College London’s ethics committee for the Institute of Psychiatry, Psychology and Neuroscience HR/DP-20/21-22060, and participant consent was collected at every stage of the TEDS data collection.

### Callous Unemotional index creation

CU traits at ages 7, 9 and 12 years old were assessed by a seven-item scale, as shown in previous work^38^. CU scores were composed of four Strengths and Difficulties Questionnaire items which cover prosocial behaviour (reverse-scored to capture anti-sociality; i.e. ‘Considerate of other people’s feelings’, ‘Helpful if someone hurt’, ‘Have at least one good friend’ and ‘Kind to younger children’) and three Antisocial Process Screening Device (APSD)^51^: CU subscale items (i.e. ‘Does not show feelings or emotions’ ‘Guilty when does something wrong (reverse-scored)’ and ‘Concerned to do well (reverse-scored)’). Each item was rated on a 3-point Likert scale (0 = Not true,1= Somewhat true and 2 = Certainly true). The CU score at age 16 was also computed from the same four SDQ items, and three items from the Inventory of Callous–Unemotional Traits (ICU)^52^ with the same item content than the APSD CU subscale, but rated on a 4-point Likert scale ranging from 0 (Not at all true) to 3 (Definitely true). When creating the composite score for age 16, to adjust score range of the ICU to the APSD, we applied a linear transformation^53^ to the ICU’s scaling changing it from 0 to 3, to 0 to 2 to match the APSD. These three item scores were multiplied by two thirds before creating the composite score in CU traits for age 16. All scores were regressed on age and sex prior to analyses in this study.

#### Polygenic score generation

To calculate polygenic scores, we used a Bayesian approach to polygenic score calculation, implemented in the software LDpred2^44^. In this method, a posterior effect size is calculated for each single SNP that is present in both the GWA study summary statistics and the target genotype sample. To calculate the posterior effect size, the original summary statistic effect size estimates are adjusted based on two factors: (a) the relative influence of a SNP given its level of LD with surrounding SNPs in the target sample (here TEDS), and (b) a prior on the effect size of each SNP. To account for LD, we set the radius to a 2 megabase window. The effect size prior depends on the SNP-heritability of the discovery (i.e., GWA study) trait and an assumption on the fraction of causal markers believed to influence the discovery trait. Using the prior, the beta effect sizes are reweighted such that the effects are spread out among the SNPs across the whole genome in proportion to the LD present among these SNPs. In the creation of the polygenic scores, summary stats were filtered to remove (1) rare SNPs (minor allele frequency < 0.005), (2) SNPs with an IMPUTE imputation quality (INFO) score < 0.9, (3) SNPs that could not be mapped to or had discrepant alleles with the reference panel and (4) otherwise low-quality variants. Thus, 6,170,305 SNPs were included analysis. Finally, all trait-associated alleles were counted (0, 1, or 2 for each SNP), weighted by the posterior SNP effect size obtained through LDpred, and summed across the genome to calculate a polygenic score for everyone in TEDS.

#### Common factor score creation

We created common factor scores using a confirmatory factor analysis with one factor per phenotype, followed by one factor per reporter, and one factor per time point for each phenotype in the R package Lavaan^50^. Supplementary Figures 1 & 2 show the structural equation models for each common factor.

## Results

### Twin models

For conduct problems, additive genetic (A) effects explained of 25-75% of the variance in conduct problems, 5-80% in ADHD symptoms and 43-58% in CU traits. Common environmental influences (C; influences which make twins more similar for a trait), were largely non-significant for conduct problems across timepoints and reporters, except for parent reported conduct problems at ages 4-12, where C estimates ranged from 5-25%. For callous unemotional traits, C explained 10-30% of the variance. When modelling ADHD symptoms, MZ twin correlations were more than double DZ twin correlations, so an ADE model (D = dominant genetic effect) was appropriate (figure 5). There was no significant contribution of D to the variance in parent-reported ADHD symptoms and so we dropped D from these models. Applying univariate twin models to common factor scores indexing shared variance across development and reporter, we found that C explained most of the variance in conduct problems (53%) and callous unemotional traits (77%), whereas A explained the majority of variance for ADHD symptoms (80%) (Figure 11). Using factors split by reporter showed that the large effect of C on conduct problems and callous unemotional traits was largely driven by parent-reported measures (Figure 12). Timepoint specific factors showed that A had the most influence on the variance across all three phenotypes (Figure 13)

#### Post-hoc polygenic analyses

Analyses of common factors indexing stability stratified by reporter or timepoint showed that these results were consistent across reporter (Figure 7a) and timepoints (Figure 8a), where significant indirect effects were found on across time within-reporter (Figure 7a; child = 30%, parent = 39% and teacher = 26% of total prediction), and across reporters within-time-points (Figure 8a; age 7 = 56%, age 9 = 35%, age 12 = 36%, age 16 = 32%, age 21 = 28% of total prediction) but not for ADHD symptoms or callous-unemotional traits (with the one exception of significant indirect genetic effect on the factor for callous-unemotional traits at age 12).

## Data Availability

All data produced in the present study are available upon reasonable request to the authors with approval from the TEDS team.

## Acknowledgements

TEDS is supported by a programme grant (MR/V012878/1) to Professor Thalia Eley from the UK Medical Research Council (previously MR/M021475/1 awarded to Professor Robert Plomin), with additional support from the US National Institutes of Health (AG046938). The authors gratefully acknowledge the ongoing contribution of the TEDS participants and their families.

The positions of J.K.B., T.A.M., Y.I.A., C.R. were funded by a Wellcome Trust Senior Research Fellowship awarded to T.A.M. (grant number 220382/Z/20/Z).

## Supplementary figures

**Supp Table 1:** Sample sizes for the whole sample (for twin models), and dizygotic (DZ) twin sample for polygenic risk score analyses, across timepoints and reporters.

| Whole sample N | Age |  |  |  |  |  |  |  |  |  |  |  |  |  |  |  |  |  |
| --- | --- | --- | --- | --- | --- | --- | --- | --- | --- | --- | --- | --- | --- | --- | --- | --- | --- | --- |
|  | 4 |  |  | 7 |  |  | 9 |  |  | 12 |  |  | 16 |  |  | 21 |  |  |
| Phenotype | Parent | Teacher | Child | Parent | Teacher | Child | Parent | Teacher | Child | Parent | Teacher | Child | Parent | Teacher | Child | Parent | Teacher | Child |
| Conduct | 15,223 | x | x | 14,864 | 12,040 | x | 6,623 | 5,606 | 6,550 | 11,382 | 9,546 | 11,362 | 9,904 | x | 9,853 | 10,352 | x | 9,189 |
| Hyperactivity | 15,216 | x | x | 14,853 | 12,031 | x | 6,619 | 5,605 | 6,550 | 11,382 | 9,546 | 11,361 | 9,890 | x | 9,854 | 10,343 | x | 9,187 |
| Callous Unemotional traits | x | x | x | 14,855 | 12,027 | x | 6,615 | 5,615 | x | 11,411 | 9,504 | x | 9,901 | x | x | x | x | x |
| DZ sample N | Age |  |  |  |  |  |  |  |  |  |  |  |  |  |  |  |  |  |
|  | 4 |  |  | 7 |  |  | 9 |  |  | 12 |  |  | 16 |  |  | 21 |  |  |
| Phenotype | Parent | Teacher | Child | Parent | Teacher | Child | Parent | Teacher | Child | Parent | Teacher | Child | Parent | Teacher | Child | Parent | Teacher | Child |
| Conduct | 5,142 | x | x | 5,266 | 4,236 | x | 2,478 | 1,940 | 2,406 | 4,524 | 3,334 | 4,480 | 4,016 | x | 3,968 | 4,220 | x | 3,042 |
| Hyperactivity | 5,148 | x | x | 5,262 | 4,232 | x | 2,478 | 1,936 | 2,406 | 4,522 | 3,330 | 4,478 | 4,008 | x | 3,970 | 4,214 | x | 3,040 |
| Callous Unemotional traits | x | x | x | 5,260 | 4,230 | x | 2,482 | 1,950 | x | 4,532 | 3,304 | x | 4,014 | x | x | x | x | x |

**Supplementary Figure 1:**
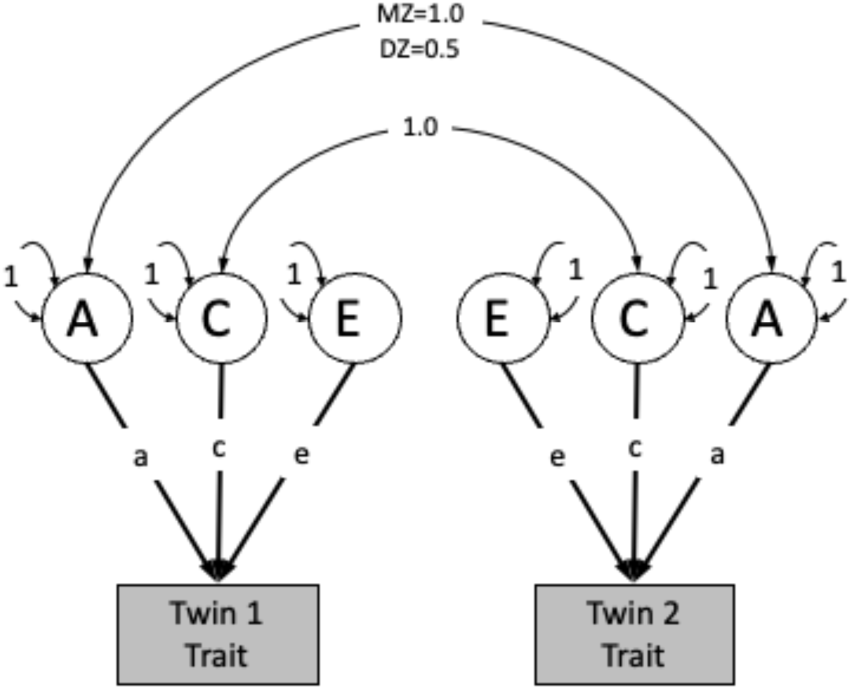
Univariate Twin model. Decomposes variance in a trait into additive genetic effects (A), common environmental influences (C) and unique environmental influences (E). Monozygotic (MZ) twins have a correlation of 1.0 for A, whereas dizygotic (DZ) twins have a correlation of 0.5. In this model, all twin pairs have a correlation of 1.0 for C.

**Supp Figure 2:**
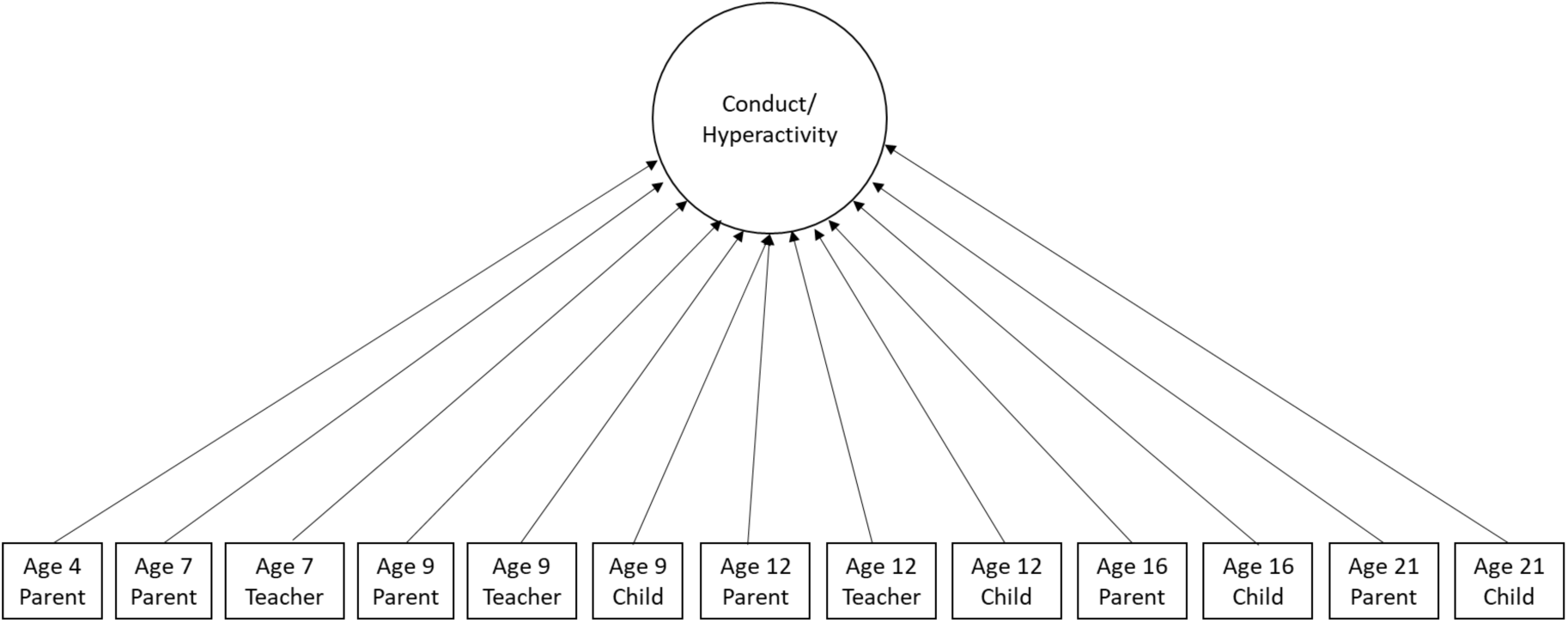
Latent factor for conduct and hyperactivity problems. Factor created from measures of externalising across all time points and reporters. We also created latent factors stratified by reporter or timepoint separately.

**Supp Figure 3:**
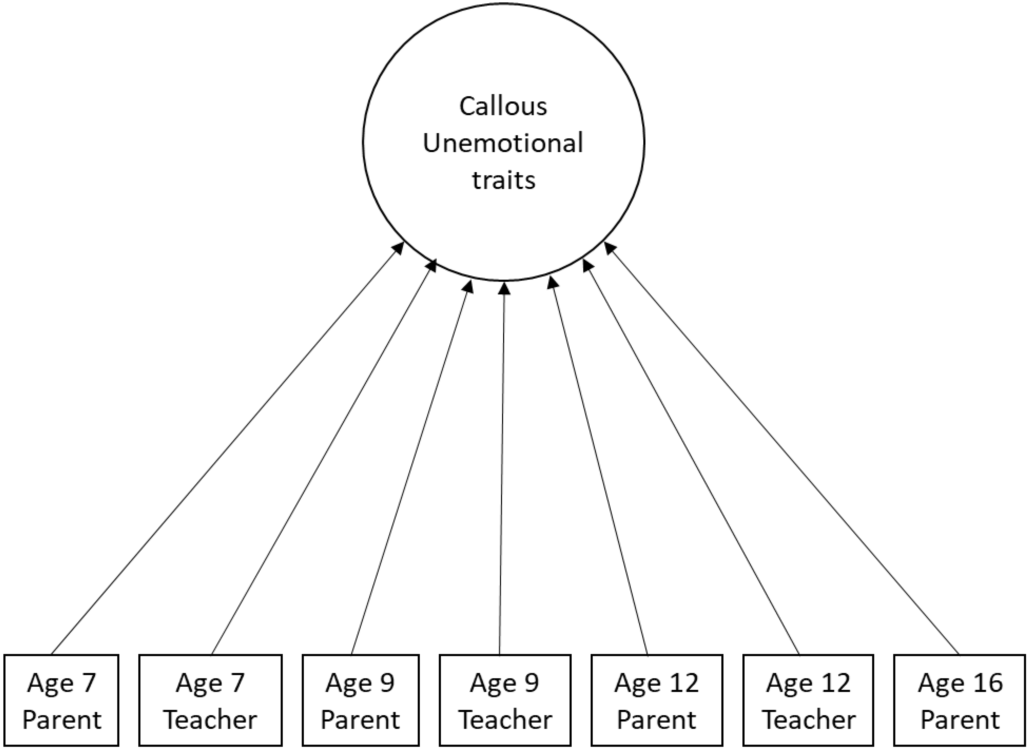
Latent factor for callous unemotional traits. Factor created from measures of externalising across all time points and reporters. We also created latent factors stratified by reporter or timepoint separately.

**Supp Figure 4:**
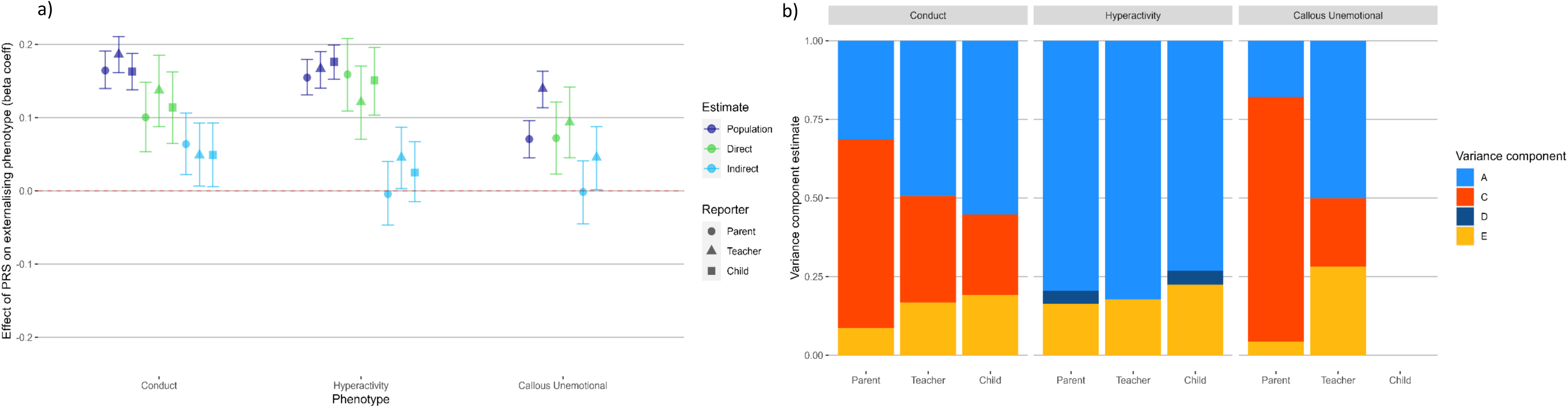
Estimating a) population-level, direct and indirect genetic effects of externalising PRS and b) univariate ACE twin models on common factor scores for each phenotype, split by reporter. Factor scores were created using common factor analysis in lavaan, extracting stability for each phenotype from measures across all timepoints, per reporter to assess whether any of the PRS prediction was driven by a specific reporter. Beta coefficient estimates of population-level prediction of externalising PRS alongside estimates of direct and indirect genetic effects, for common factor scores created for conduct problems, ADHD symptoms and callous-unemotional traits, Estimates of the contribution of additive genetic effects (A), common environmental influences (C) and unique environmental influences (E) in the variance of each factor, per reporter to assess whether estimates of influences on the variance per common factor differed across reporters.

**Supp Figure 5:**
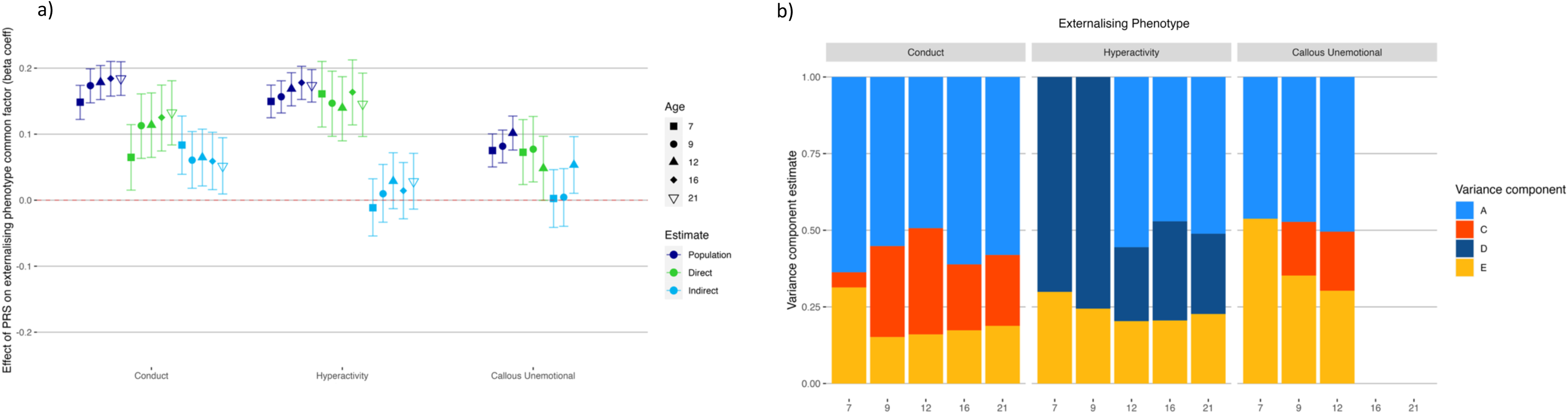
Estimating a) population-level, direct and indirect genetic effects of externalising PRS and b) univariate ACE twin models of common factor scores for each externalising phenotype, split by age. Factor scores were created using common factor analysis in lavaan, extracting stability for each phenotype from measures across all reporters, to assess whether any of the PRS prediction was driven by externalising symptoms at a specific age during development. Beta coefficient estimates of population-level prediction of externalising PRS alongside estimates of direct and indirect genetic effects, for common factor scores created for each externalising phenotype. Estimates of the contribution of additive genetic effects (A), common environmental influences (C), dominant genetic effects (D) and unique environmental influences (E) in the variance of each factor.

**Supp Figure 6:**
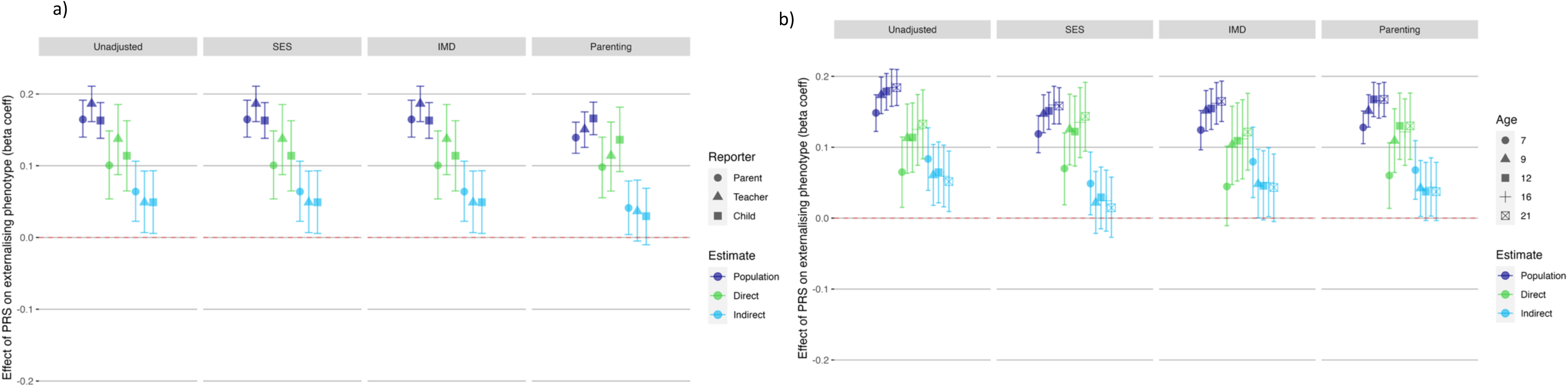
Investigating impact of socioeconomic status indices or parenting variables on direct and indirect genetic effects of externalising PRS on a) within-reporter common factor and b) within-timepoint common factor for conduct problems. Socioeconomic status (SES) was measured at first contact and comprises measures of parent employment, education, and age of mother on first birth. The Indices of Multiple Deprivation (IMD) decile score uses census data matched with participants post codes, giving a broader measure of wider environmental factors such as local levels of employment and education, crime rates, barriers to housing and living environment quality. The parenting analyses used a latent factor created from ‘Parental Feelings’ and ‘Parental Discipline’ rated by parent at ages 4, 7, 9 and 12.

## References

1. Erskine, H. E. et al. Long-Term Outcomes of Attention-Deficit/Hyperactivity Disorder and Conduct Disorder: A Systematic Review and Meta-Analysis. J. Am. Acad. Child Adolesc. Psychiatry 55, 841–850 (2016).

2. Wertz, J. et al. From Childhood Conduct Problems to Poor Functioning at Age 18 Years: Examining Explanations in a Longitudinal Cohort Study. J. Am. Acad. Child Adolesc. Psychiatry 57, 54–60.e4 (2018).

3. De Brito, S. A. et al. Psychopathy. Nat. Rev. Dis. Primer 7, 1–21 (2021).

4. Thøgersen, D. M., Elmose, M., Viding, E., McCrory, E. & Bjørnebekk, G. Behavioral Improvements but Limited Change in Callous-Unemotional Traits in Adolescents Treated for Conduct Problems. J. Child Fam. Stud. 31, 3342–3358 (2022).

5. Dhanani, S. et al. A systematic review of the heritability of specific psychopathic traits using Hare’s two-factor model of psychopathy. CNS Spectr. 23, 29–38 (2018).

6. Moore, A. A., Blair, R. J., Hettema, J. M. & Roberson-Nay, R. The Genetic Underpinnings of Callous-Unemotional Traits: A Systematic Research Review. Neurosci. Biobehav. Rev. 100, 85–97 (2019).

7. Saunders, M. C. et al. The Associations between Callous-unemotional Traits and Symptoms of Conduct Problems, Hyperactivity and Emotional Problems: A Study of Adolescent Twins Screened for Neurodevelopmental Problems. J. Abnorm. Child Psychol. 47, 447–457 (2019).

8. Karlsson Linnér, R., et al. Multivariate analysis of 1.5 million people identifies genetic associations with traits related to self-regulation and addiction. Nat. Neurosci. 24, 1367–1376 (2021).

9. Wang, F. L., Bountress, K. E., Lemery-Chalfant, K., Wilson, M. N. & Shaw, D. S. A Polygenic Risk Score Enhances Risk Prediction for Adolescents’ Antisocial Behavior over the Combined Effect of 22 Extra-familial, Familial, and Individual Risk Factors in the Context of the Family Check-Up. Prev. Sci. 24, 739–751 (2023).

10. Anderson, R. E. et al. Pathways to Pain: Racial Discrimination and Relations Between Parental Functioning and Child Psychosocial Well-Being. J. BLACK Psychol. 41, 491–512 (2015).

11. Adeyemo, A. et al. Responsible use of polygenic risk scores in the clinic: potential benefits, risks and gaps. Nat. Med. 27, 1876–1884 (2021).

12. Burgess, S., Foley, C. N. & Zuber, V. Inferring Causal Relationships Between Risk Factors and Outcomes from Genome-Wide Association Study Data. Annu. Rev. Genomics Hum. Genet. 19, 303–327 (2018).

13. Uddin, M. J., Hjorthøj, C., Ahammed, T., Nordentoft, M. & Ekstrøm, C. T. The use of polygenic risk scores as a covariate in psychological studies. Methods Psychol. 7, 100099 (2022).

14. Cheesman, R., Ayorech, Z., Eilertsen, E. M. & Ystrom, E. Why we need families in genomic research on developmental psychopathology. JCPP Adv. 3, e12138 (2023).

15. Demange, P. A. et al. Estimating effects of parents’ cognitive and non-cognitive skills on offspring education using polygenic scores. Nat. Commun. 13, 4801 (2022).

16. Eilertsen, E. M. et al. On the importance of parenting in externalizing disorders: an evaluation of indirect genetic effects in families. J. Child Psychol. Psychiatry 63, 1186–1195 (2022).

17. Kong, A. et al. The nature of nurture: Effects of parental genotypes. Science 359, 424–428 (2018).

18. McAdams, T. A., Cheesman, R. & Ahmadzadeh, Y. I. Annual Research Review: Towards a deeper understanding of nature and nurture: combining family-based quasi-experimental methods with genomic data. J. Child Psychol. Psychiatry (2022) doi:10.1111/jcpp.13720.

19. Selzam, S. et al. Comparing Within- and Between-Family Polygenic Score Prediction. Am. J. Hum. Genet. 105, 351–363 (2019).

20. Wang, B. et al. Robust genetic nurture effects on education: A systematic review and meta-analysis based on 38,654 families across 8 cohorts. Am. J. Hum. Genet. 108, 1780–1791 (2021).

21. Hicks, B. M., Krueger, R. F., Iacono, W. G., McGue, M. & Patrick, C. J. Family transmission and heritability of externalizing disorders: A Twin-Family Study. Arch. Gen. Psychiatry 61, 922–928 (2004).

22. Rhule, D. & McMahon, R. Problem Behavior and Romantic Relationships: Assortative Mating, Behavior Contagion, and Desistance. Clin. Child Fam. Psychol. Rev. 10, 53–100 (2007).

23. Patalay, P., Fink, E., Fonagy, P. & Deighton, J. Unpacking the associations between heterogeneous externalising symptom development and academic attainment in middle childhood. Eur. Child Adolesc. Psychiatry 25, 493–500 (2016).

24. Schmengler, H. et al. Educational level, attention problems, and externalizing behaviour in adolescence and early adulthood: the role of social causation and health-related selection—the TRAILS study. Eur. Child Adolesc. Psychiatry 32, 809–824 (2023).

25. Morris, T. T., Davies, N. M., Hemani, G. & Smith, G. D. Population phenomena inflate genetic associations of complex social traits. Sci. Adv. 6, eaay0328 (2020).

26. Pingault, J.-B. et al. Identifying intergenerational risk factors for ADHD symptoms using polygenic scores in the Norwegian Mother, Father and Child Cohort. 2021.02.16.21251737 Preprint at 10.1101/2021.02.16.21251737 (2021).

27. de Zeeuw, E. L. et al. Intergenerational Transmission of Education and ADHD: Effects of Parental Genotypes. Behav. Genet. 50, 221–232 (2020).

28. Demontis, D. et al. Discovery of the first genome-wide significant risk loci for attention deficit/hyperactivity disorder. Nat. Genet. 51, 63–75 (2019).

29. Plomin, R. & von Stumm, S. The new genetics of intelligence. Nat. Rev. Genet. 19, 148–159 (2018).

30. Rijsdijk, F. V. & Sham, P. C. Analytic approaches to twin data using structural equation models. Brief. Bioinform. 3, 119–133 (2002).

31. Cheesman, R. et al. How important are parents in the development of child anxiety and depression? A genomic analysis of parent-offspring trios in the Norwegian Mother Father and Child Cohort Study (MoBa). medRxiv 1–11 (2020) doi:10.1101/2020.04.14.20064782.

32. Flom, M. & Saudino, K. J. Do Genetic Factors Explain the Links Between Callous-Unemotional, Attention Hyperactivity and Oppositional Defiant Problems in Toddlers? J. Abnorm. Child Psychol. 46, 1217–1228 (2018).

33. Larsson, H., Andershed, H. & Lichtenstein, P. A genetic factor explains most of the variation in the psychopathic personality. J. Abnorm. Psychol. 115, 221–230 (2006).

34. Viding, E., Frick, P. J. & Plomin, R. Aetiology of the relationship between callous-unemotional traits and conduct problems in childhood. Br. J. Psychiatry 190, s33–s38 (2007).

35. Hannigan, L. J., Walaker, N., Waszczuk, M. A., McAdams, T. A. & Eley, T. C. Aetiological Influences on Stability and Change in Emotional and Behavioural Problems across Development: A Systematic Review. Psychopathol. Rev. a4, 52–108 (2017).

36. Pingault, J.-B., Rijsdijk, F., Zheng, Y., Plomin, R. & Viding, E. Developmentally dynamic genome: Evidence of genetic influences on increases and decreases in conduct problems from early childhood to adolescence. Sci. Rep. 5, 10053 (2015).

37. Pingault, J.-B. et al. Genetic and Environmental Influences on the Developmental Course of Attention-Deficit/Hyperactivity Disorder Symptoms From Childhood to Adolescence. JAMA Psychiatry 72, 651–658 (2015).

38. Takahashi, Y., Pease, C. R., Pingault, J. B. & Viding, E. Genetic and environmental influences on the developmental trajectory of callous-unemotional traits from childhood to adolescence. J. Child Psychol. Psychiatry 62, 414–423 (2021).

39. Pingault, J.-B. et al. Genetic and Environmental Influences on the Developmental Course of Attention-Deficit/Hyperactivity Disorder Symptoms From Childhood to Adolescence. JAMA Psychiatry 72, 651–658 (2015).

40. Rimfeld, K. et al. Twins Early Development Study: A Genetically Sensitive Investigation into Behavioral and Cognitive Development from Infancy to Emerging Adulthood. Twin Res. Hum. Genet. 22, 508–513 (2019).

41. Lockhart, C. et al. Twins Early Development Study (TEDS): A genetically sensitive investigation of mental health outcomes in the mid-twenties. JCPP Adv. (2023) doi:10.1002/jcv2.12154.

42. Goodman, R. Psychometric properties of the strengths and difficulties questionnaire. Journal of the American Academy of Child and Adolescent Psychiatry vol. 40 1337–1345 (2001).

43. Haider, Z. F. & von Stumm, S. Predicting educational and social–emotional outcomes in emerging adulthood from intelligence, personality, and socioeconomic status. J. Pers. Soc. Psychol. 123, 1386–1406 (2022).

44. Privé, F., Arbel, J. & Vilhjálmsson, B. J. LDpred2: better, faster, stronger. Bioinforma. Oxf. Engl. 36, 5424–5431 (2021).

45. R Core Team. R: A Language and Environment for Statistical Computing R. (2023).

46. Rietveld, M. J. H., Posthuma, D., Dolan, C. V. & Boomsma, D. I. ADHD: Sibling Interaction or Dominance: An Evaluation of Statistical Power. Behav. Genet. 33, 247–255 (2003).

47. Boker, S. M., et al. OpenMx: Extended Structural Equation Modelling. (2023).

48. Merwood, A. et al. Different heritabilities but shared etiological influences for parent, teacher and self-ratings of ADHD symptoms: an adolescent twin study. Psychol. Med. 43, 1973–1984 (2013).

49. Cheesman, R. et al. Extracting stability increases the SNP heritability of emotional problems in young people. Transl. Psychiatry 8, 223 (2018).

50. Rosseel, Y. lavaan: An R Package for Structural Equation Modeling. J. Stat. Softw. 48, 1–36 (2012).

51. Frick, P. J. & Hare, R. D. Antisocial process screening device. Eur. J. Psychol. Assess. (2001).

52. Frick, P. J. Inventory of callous–unemotional traits. PLoS One (2004).

53. Colman, A. M., Norris, C. E. & Preston, C. C. Comparing rating scales of different lengths: Equivalence of scores from 5-point and 7-point scales. Psychol. Rep. 80, 355–362 (1997).

